# Unsupervised Ensemble Learning for Efficient Integration of Pre-trained Polygenic Risk Scores

**DOI:** 10.1101/2025.01.06.25320058

**Authors:** Chenyin Gao, Justin D. Tubbs, Yi Han, Min Guo, Sijia Li, Erica Ma, Dailin Luo, Jordan W. Smoller, Phil H. Lee, Rui Duan

## Abstract

The growing availability of pre-trained polygenic risk score (PRS) models has enabled their integration into real-world applications, reducing the need for extensive data labeling, training, and calibration. However, selecting the most suitable PRS model for a specific target population remains challenging, due to issues such as limited transferability, data heterogeneity, and the scarcity of observed phenotype in real-world settings. Ensemble learning offers a promising avenue to enhance the predictive accuracy of genetic risk assessments, but most existing methods often rely on observed phenotype data or additional genome-wide association studies (GWAS) from the target population to optimize ensemble weights, limiting their utility in real-time implementation. Here, we present the <u>UN</u>supervised en<u>Semble</u> <u>PRS</u> (*UNSemblePRS*), an unsupervised ensemble learning framework, that combines pre-trained PRS models without requiring phenotype data or summaries from the target population. Unlike traditional supervised approaches, UNSemblePRS aggregates models based on prediction concordance across a curated subset of candidate PRS models. We evaluated UNSemblePRS using both continuous and binary traits in the All of Us database, demonstrating its scalability and robust performance across diverse populations. These results underscore UNSemblePRS as an accessible tool for integrating PRS models into real-world contexts, offering broad applicability as the availability of PRS models continues to expand.

## 1 Introduction

Polygenic risk score (PRS) has emerged as a powerful tool to predict an individual’s genetic predisposition to specific phenotypes. Typically, PRSs aggregate hundreds to millions of genetic variants identified in genome-wide association studies (GWAS) through a weighted sum of allele doses, each multiplied by its respective effect size (Khera et al., 2018; Kullo et al., 2022). Over the past decades, numerous PRS methods, such as LDpred (Vilhjálmsson et al., 2015), LDpred2 (Privé et al., 2020), lassosum (Mak et al., 2017), and PRS-CS (Ge et al., 2019), have been developed to more accurately quantify genetic predisposition for a variety of clinically relevant measures (e.g., body mass index (BMI) (Khera et al., 2019)) and diseases (e.g., breast cancer (BC) (Mavaddat et al., 2019)). For many clinical use cases, PRSs are becoming increasingly important for risk stratification and early identification of high-risk individuals.

With the growing emphasis on open science, researchers are progressively making their PRS models publicly accessible. Efforts to support this movement include the development of open-source repositories like the PGS Catalog (https://www.pgscatalog.org), which archives and documents publicly available PRS models. In just several years, over 5000 models spanning more than 650 traits have been shared, with this number continuing to grow rapidly. For widely studied traits such as height, Body Mass Index (BMI), and Breast Cancer (BC), more than 100 pre-trained models are available. These models, developed by various research groups, utilize diverse statistical and machine learning techniques applied to a range of datasets. The availability of these models promotes transparency, reproducibility, and collaboration in genetic research, creating new opportunities that leverages model-sharing and the integration of diverse data sources (Lambert et al., 2024b).

Despite recent advancements, from an implementation perspective, practitioners aiming to apply PRS models for risk stratification or other tasks requiring the quantification of genetic risk for specific traits often encounter uncertainty regarding best practices for leveraging pre-trained models in their own practice. This uncertainty arises from several factors. First, while a wide range of methods for training PRS models is available, there is no strong evidence that any one method consistently outperforms others (Ma and Zhou, 2021). Furthermore, the performance of PRS models is known to vary significantly across different biobanks and populations (Wojcik et al., 2019; Duncan et al., 2019; Fatumo et al., 2022), due to potential differences in contextual factors such as age, sex, and health status. Another significant challenge is that most GWAS studies predominantly include individuals of European (EUR) ancestry, leading to PRS models derived from these studies often performing poorly in non-EUR populations (Ding et al., 2022; Hou et al., 2024).

These uncertainties make it challenging to directly select the most suitable pre-trained PRS models for a given target population in a real-world implementation. To address this limitation, increasing efforts have been made to develop new PRS models by leveraging ancestry-specific GWAS summary statistics or utilizing multi-population GWAS meta-analyses that incorporate cross-population data to develop more robust PRS models with improved transferability across diverse populations (Ruan et al., 2022; Xiao et al., 2022). While retraining or calibrating PRS models for target populations can improve predictive accuracy, this approach is often not feasible due to the lack of complete or available GWAS data and linkage disequilibrium (LD) matrices for many non-EUR populations (Wray et al., 2007; Martin et al., 2017). Additionally, developing new PRS models requires substantial computational resources, stringent quality control processes, and multidisciplinary expertise in data pre-processing. These demands significantly hinder the practicality of widespread PRS model development and application.

As pre-trained models become more widely accessible, the development of effective methods for combining them has gained increasing importance, enabling researchers to leverage the strengths of individual models for improved overall performance. Various ensemble techniques have been designed to aggregate multiple PRS models tailored to target populations. Examples include multi-PRS (Albiñana et al., 2023), PRSmix (Truong et al., 2024), and PROSPER (Zhang et al., 2024), which employ ensemble learning strategies such as lasso, ridge, and SuperLearner to combine PRS models and optimize predictive power for specific populations or phenotypes (Márquez-Luna et al., 2017; Lloyd-Jones et al., 2019; Zhao et al., 2024).

However, training these ensemble models often requires phenotype data or additional GWAS from the target population, which may not always be readily available. For instance, evaluating genetic risk scores for a particular disease in a relatively young population can be challenging, as most individuals may not yet have experienced disease onset. Additionally, accurately defining the phenotype of interest for supervised ensemble training from large datasets, such as electronic health records, can be both costly and labor-intensive, often necessitating specialized clinical expertise (Lobach and Detmer, 2007). These challenges present significant barriers to the widespread application of supervised PRS ensemble methods.

In this paper, we introduce an easy-to-implement UNsupervised enSemble PRS (*UNSemblePRS*) framework that leverages pre-trained PRS models without requiring phenotype data from the target population. Unlike traditional supervised ensemble learning approaches, which derive weights by optimizing predictive power for the target trait, our unsupervised method aggregates models by leveraging prediction concordance across multiple PRS models. This approach mitigates unwanted prediction variability across models, resulting in improved prediction accuracy. UNSemblePRS enables users to specify a collection of candidate models tailored to their target population of interest and generates an aggregated PRS score that delivers stable performance. It is robust in scenarios where some candidate models are of lower quality or perform poorly in the target population. Furthermore, UNSemblePRS is highly scalable and can be flexibly adjusted to accommodate real-time population shifts, making it an effective and adaptable solution for diverse research applications. Integrated with open-source resources such as the PGS Catalog, UNSemblePRS provides a valuable tool for the research community, promoting accessibility, scalability, and reproducibility in genetic risk prediction. Using data from the All of Us (AoU) Research Hub, we evaluate the performance of UNSemblePRS, benchmarking it against the best-performing single PRS predictions across multiple phenotypes. Our results demonstrate its ability to provide high-quality genetic predictions across diverse traits. In addition, we assess the transferability and interpretability of UNSemblePRS, highlighting its strengths, limitations, and potential for future applications as more PRS models become publicly available.

## 2 Results

### 2.1 Overview of Methods and Study Design

#### The UNSemblePRS method

The UNSemblePRS method integrates multiple pre-trained PRS models to generate aggregated risk scores for a target cohort of *n* individuals with available genotypic data. For a specific trait of interest, the process begins by selecting a set of candidate PRS models, which may have been trained on diverse populations, datasets, or using different PRS methodologies. In this work, we utilize models from the PGS catalog; however, users have the flexibility to curate candidate models based on their available resources and preferences. For each candidate model, PRS scores are computed for all individuals in the target cohort. With *p* candidate models, this step produces an *n × p* matrix, where the entry at position (*i, j*) corresponds to the PRS score for the *i*-th individual derived from the *j*-th model. This matrix serves as the basis for subsequent aggregation steps, enabling the generation of robust and comprehensive risk predictions.

The core of UNSemblePRS lies in assigning appropriate weights to enhance predictive power and ensure robustness across different environments. From an implementation standpoint, we assume that the trait of interest is not observed in the target cohort, which requires assigning weights in an unsupervised manner.

UNSemblePRS builds upon prior work on unsupervised model aggregation. Specifically, Parisi et al. (2014) introduced a meta-classifier that integrates a set of binary classifiers under the assumption of conditional independence between classifiers. Ma et al. (2023) extended this idea to integrate multiple dimension reduction algorithms, assuming the output of each algorithm can be expressed as a signal plus homoskedastic Gaussian noise. Coombes et al. (2020) proposed using principal component analysis (PCA) to refine tuning parameter choices in PRS models. All the aforementioned methods share a common strategy: applying spectral decomposition to a concordance matrix derived from the outputs of all classifiers or algorithms and obtaining a consensus output through a weighted average of the models, with the weights determined by the eigenvectors.

UNSemblePRS employs sparse kernel PCA on the standardized PRS matrix of dimension *n×p* to capture relationships among the PRS models (Figure 1). The standardized PRS matrix is constructed by centering and normalizing the PRS scores from each candidate model to have a mean of zero and a variance of one. The kernel function, which defines the similarity between PRS scores, maps the PRS matrix into a higher-dimensional feature space. In our work, we choose the kernel function as a polynomial function of degree three, where its explicit form can be found in the Online Method; results based on different kernel functions are included in the Supplementary Material for sensitivity analysis. This mapping enables the capture of more flexible and nonlinear relationships among the PRS scores. Within this feature space, sparse PCA is applied to identify a set of sparse eigenvectors that can reconstruct the features as accurately as possible, where the non-zero entries in the eigenvector indicate the selection of PRS models. Finally, the aggregation weights for combining the selected PRS scores into the final aggregated PRS are derived from the eigenvector generated by non-sparse kernel PCA separately for each ancestry population in the target cohort.

**Figure 1:**
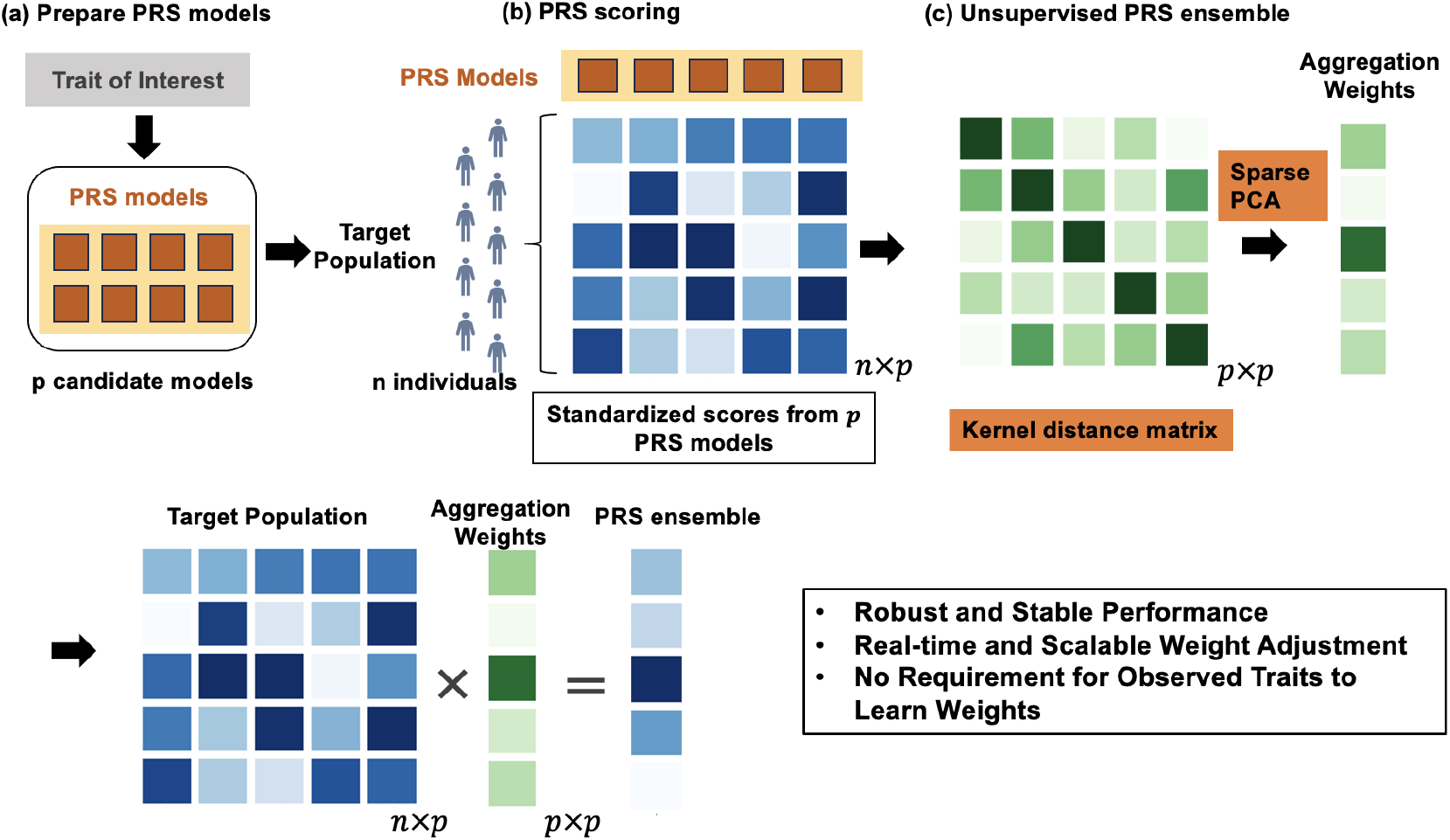
Workflow of UnsemblePRS, including 1) preparing PRS models, 2) applying the PRS models on the target population 3) performing UnsemblePRS to obtain the final aggregated score.

The rationale behind this approach is that mapping the scores into a non-linear space using the kernel product allows the leading eigenvector, which captures the largest portion of variation among the models, to effectively reflect the relative importance of the PRS models. In addition, the sparsity assumption helps mitigate issues arising from models with redundant information, low-quality training data, or errors introduced during uploading or downloading processes. These considerations are particularly relevant for open-source models available in the PGS catalog. First, many models are trained on the same GWAS data sources, leading to redundant information. Second, some models are constructed using only significant single-nucleotide polymorphisms (SNPs) from underpowered studies, resulting in scores that may be nearly non-predictive. Third, as the PGS catalog allows users to upload models, human errors, such as documenting the wrong trait or misaligning PGS weights, can occur. As demonstrated in our empirical study, applying a sparsity threshold to exclude less important models improves predictive performance. In UNSemblePRS, we set a default threshold of 50%, excluding PRS models assigned weights in the lower half. However, this threshold is adjustable based on users’ prior knowledge of redundancy or potential adversarial models among the candidates. A detailed mathematical formulation and step-by-step procedures are provided in the Online Method.

#### Evaluation of UNSemblePRS using All of Us Data

The performance of UNSemblePRS is evaluated using data from the AoU Research Hub (All of Us Research Program, 2019), a large and diverse U.S.-based cohort study supporting biomedical research to improve health outcomes. We assess the performance of UNSemblePRS on five continuous traits and five binary traits. The continuous traits include height, BMI, high-density lipoprotein (HDL) cholesterol, low-density lipoprotein (LDL) cholesterol, and diastolic blood pressure (DBP). The binary traits include BC, inflammatory bowel disease (IBD), chronic kidney disease (CKD), atrial fibrillation (AF), and coronary artery disease (CAD). The Experimental Factor Ontology (EFO) IDs of these traits can be found in the Supplementary Table A1. We downloaded all PRS models available in the PGS catalog as of May 2024 for each trait, corresponding to each trait based on their EFO IDs. The PRS models that either do not contain chromosome and position information or have no overlapping SNPs with individuals from the AoU database are excluded from our analysis. A total of 85 models are scored for BMI, 93 for height, 90 for HDL cholesterol, 104 for LDL cholesterol, and 51 for DBP. For the binary traits, we scored 135 models for BC, 20 for IBD, 20 for CKD, 37 for AF, and 67 for CAD.

To evaluate the proposed method, a total of 285,269 individuals aged 21 and older from the AoU database with available genotypic data were extracted. The target cohort for evaluation was randomly selected from a subset of participants with observed phenotypes for the trait of interest. Detailed sample size information for each phenotype is provided in Supplementary Table A1.The target cohort size is set to 10000, representing the scale of a moderately sized study in real-world practice where PRS applications are needed for various downstream tasks, such as developing a risk prediction model or accounting for genetic risk in comparative effectiveness studies. In addition to the genotype and phenotype data, we also extracted a set of covariates including age, sex, and calculated the 30 genetic principal components (PCs) for the target cohort (Price et al., 2006). Leveraging the diversity of the AoU data, we are able to assess the performance of UNSemblePRS in both the overall population and in different subpopulations defined by ancestral groups and other demographic factors. More details regarding the quality control of the genotypic data, ancestry annotation, and extraction of each trait can be found in the Online Method.

We compare UNSemblePRS to several alternative methods for unsupervised model aggregation. These include (1) the average of all standardized PRS models, which assigns equal importance to each candidate model (denoted as “Avg”), and (2) the average of PRSs weighted by the size of the relevant GWAS studies (denoted as “SSAvg”). Additionally, we consider using (3) the PRS derived from the largest GWAS study (denoted as “MaxPRS”), motivated by emerging evidence that the size of the GWAS study can significantly impact the predictive power of PRS models (Márquez-Luna et al., 2017; Grinde et al., 2019; Weissbrod et al., 2022; Lehmann et al., 2023). We also consider (4) the approach involves deriving weights using PCA applied to the correlation matrix of the candidate PRS models (denoted as “PCA”), as proposed by Coombes et al. (2020) for combining PRS models with different tuning parameters using the clumping and thresholding method. Beyond these unsupervised aggregation methods, we also benchmark UNSemblePRS against (5) the best-performing candidate model selected based on observed traits (denoted as “best”). However, it is important to note that in real-world scenarios where the traits are unobserved, identifying the best model is not feasible.

For the predictive performance assessment with continuous traits, we report the increase in explained variance on the linear scale, measured as the incremental *R*^2^. This is estimated by the difference between the *R*^2^ of the full linear model, which includes PRS and covariates, and the reduced linear model with only covariates. For binary traits, we provide the adjusted area under the receiver operating characteristic curve (AUC), accounting for the effects of covariates. In addition to assessing the predictive performance of UNSemblePRS, we examined its transferability across the context-specific populations, the impact of sparsity thresholds on model performance, and the interpretability of its weights compared to relevant benchmark methods. Further details of the implementation are provided in the Online Method section.

### 2.2 Key findings

**Unsupervised PRS ensemble demonstrates comparable performance to the best PRS across multiple traits and diverse ancestry populations** Figure 2 illustrates the predictive performance of the compared methods for both quantitative and binary traits. Meanwhile, we also calculate the difference in predictive accuracy for each unsupervised method relative to the best single PRS model, averaged across all continuous or binary traits (see the heatmaps on the left of Figure 2). These evaluations are conducted on the entire target cohort (ALL) and stratified by the ancestry annotation; additional evaluation on the performance stratified by the age and sex are provided in the Supplementary Figures A1 and A2.

**Figure 2:**
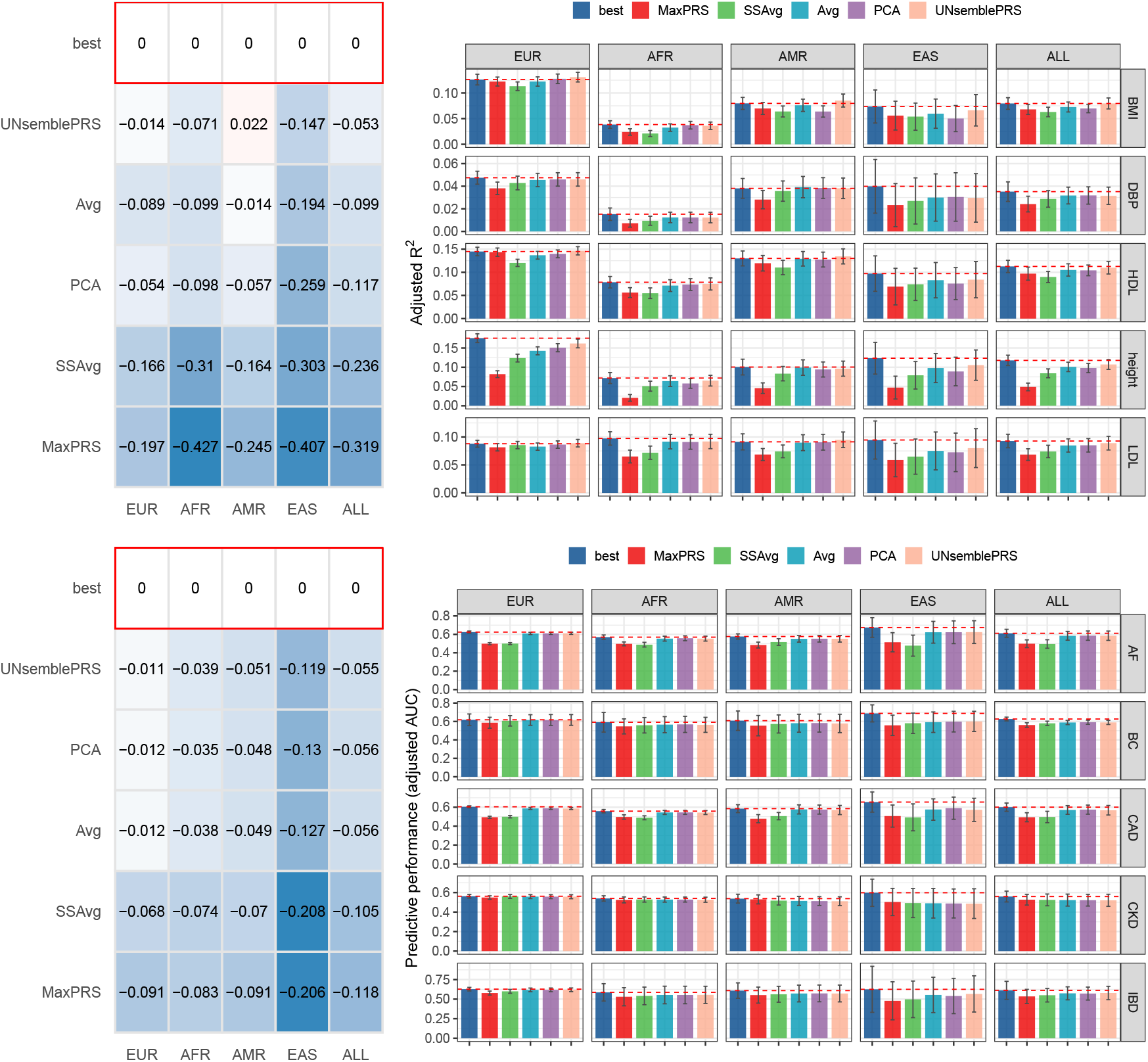
**(Left)** Relative differences in predictive performance compared to the best single PRS model, evaluated across 100 random samples (N = 10,000) and averaged over continuous and binary traits in the overall target cohort and stratified by genetic ancestry. **(Right)** Predictive performance for continuous and binary traits in the overall target cohort and across ancestry groups. Error bars represent *±*1 standard deviation computed from the random samples. Dashed lines denote the performance of the best single PRS model.

For continuous traits, UNSemblePRS achieves predictive accuracy comparable to the best single PRS model across all ancestry populations. On average, its performance difference is around 5% relative to the best model in the overall population, less than 3% in EUR and American (AMR) populations, 7% in African-ancestry population (AFR) and 14% in East Asian population (EAS). Notably, UNSemblePRS significantly outperforms other unsupervised methods for aggregating models.On average across continuous traits, UNSemblePRS exhibits the smallest relative decrease in performance compared to the best single model, being 46.9% smaller than the unweighted average and 55.1% smaller than PCA. Methods that aggregate PRS models using GWAS sample sizes or selected based on the largest GWAS sample size perform worse than other approaches, suggesting that larger GWAS sample sizes do not directly translate into better predictive performance for PRS models. In addition, UNSemblePRS remains the most effective unsupervised method for predicting binary traits. However, the improvement is less obvious compared to the performance for continuous traits. Predicting binary traits poses challenges, as many candidate models from the PGS catalog exhibit relatively low predictive power in the AoU database. Consequently, this limited model performance leads to similar weights being assigned to each candidate model across multiple ancestry populations, causing the performance more similar to the average method.

#### UNSemblePRS assigns higher weights to models with better predictive performance

We show that the weights learned by UNSemblePRS demonstrate stronger alignment with the true predictive performance of each candidate model. For each population, we assess the predictive performance of each model using the observed traits and regress it against the weights assigned by PCA and UNSemblePRS after proper scaling.

Figure 3 presents scatter plots and fitted curves for all the candidate PRS models across the considered traits. Both weights and true predictive performance are normalized to have zero mean and unit variance for each set of trait-specific candidate PRS models. The fitted *R*^2^ values are used to evaluate alignment or concordance between the actual model performance and the assigned weights. For continuous traits, the concordance can reach as high as *R*^2^ = 0.79, indicating that models with higher predictive performance are consistently assigned larger weights. In contrast, for the PCA method, the concordance can be as low as 0.01, suggesting that higher weights may be assigned to models with relatively low performance. For binary traits, the weights assigned by UNSemblePRS show weaker concordance with true predictive performance, especially in non-EUR populations, although they still outperform PCA weights in alignment. This issue likely arises because most PRS models for binary traits in the PGS catalog are trained on EUR GWAS samples, resulting in poor and indistinguishable performance in non-EUR populations. These findings highlight UNSemblePRS’s ability to effectively estimate the relative performance of each candidate model and rank them accordingly for both continuous and binary traits.

**Figure 3:**
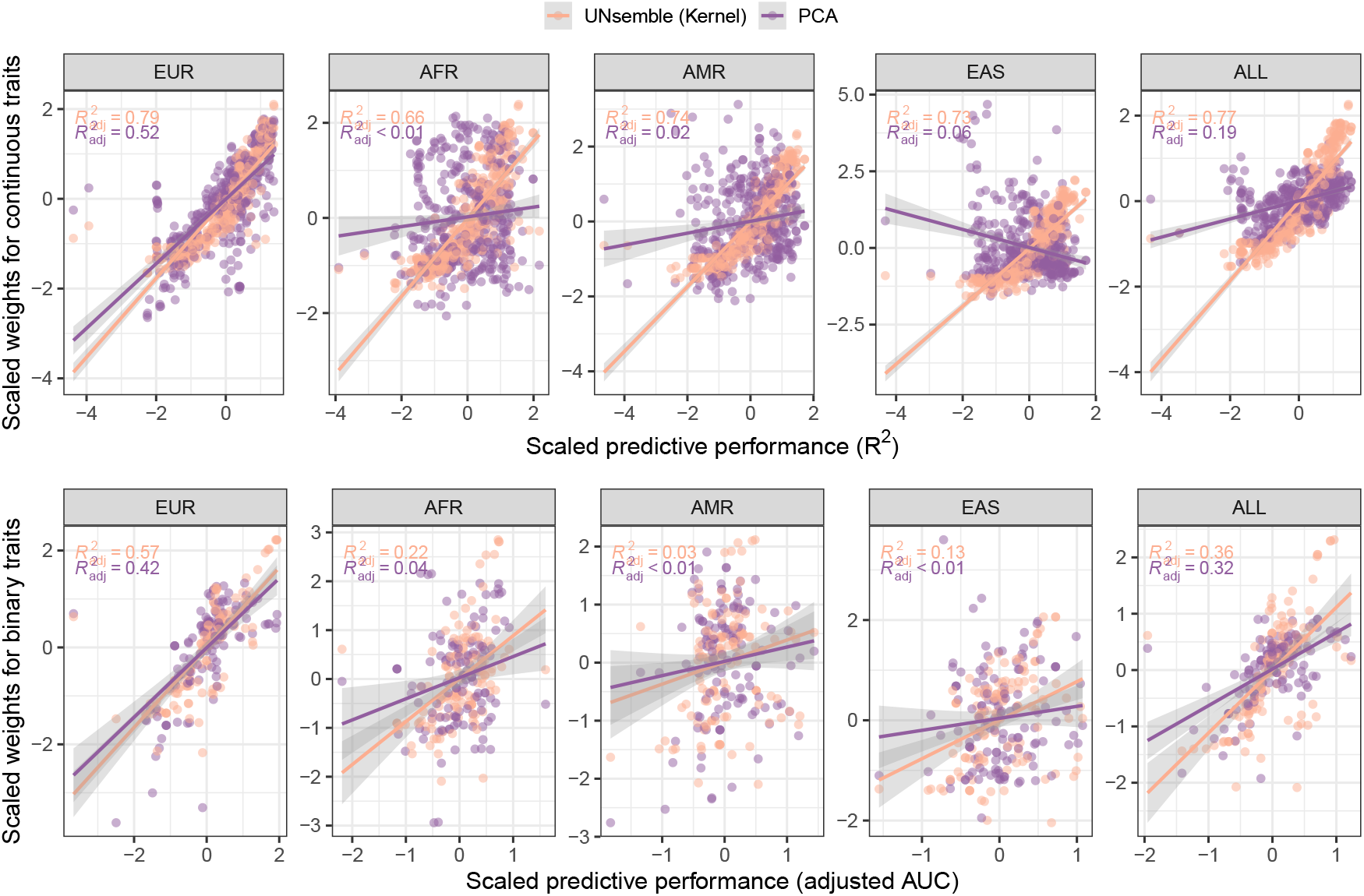
The scaled aggregation weights assigned to each candidate PRS model by the kernel and standard PCA methods, compared to the corresponding scaled predictive performance (*R*^2^ or adjusted AUC) across multiple ancestry populations and the overall cohort. Each point represents a PRS model from the PGS Catalog.

#### UNSemblePRS shows better transferability than the best model

The best PRS model, identified using the observed traits, often varies across populations. This variation can lead to transferability challenges, as the best model for one dataset may perform poorly in another population. To illustrate this, we utilize word clouds (Figure 4) to show the frequency at which each trait-specific PRS model is identified as the best model across 100 independent target cohorts within each ancestry population. The size of each PRS model’s name in the word cloud is proportional to the number of times it is selected as the best model, with models fewer than twice excluded for clarity.

**Figure 4:**
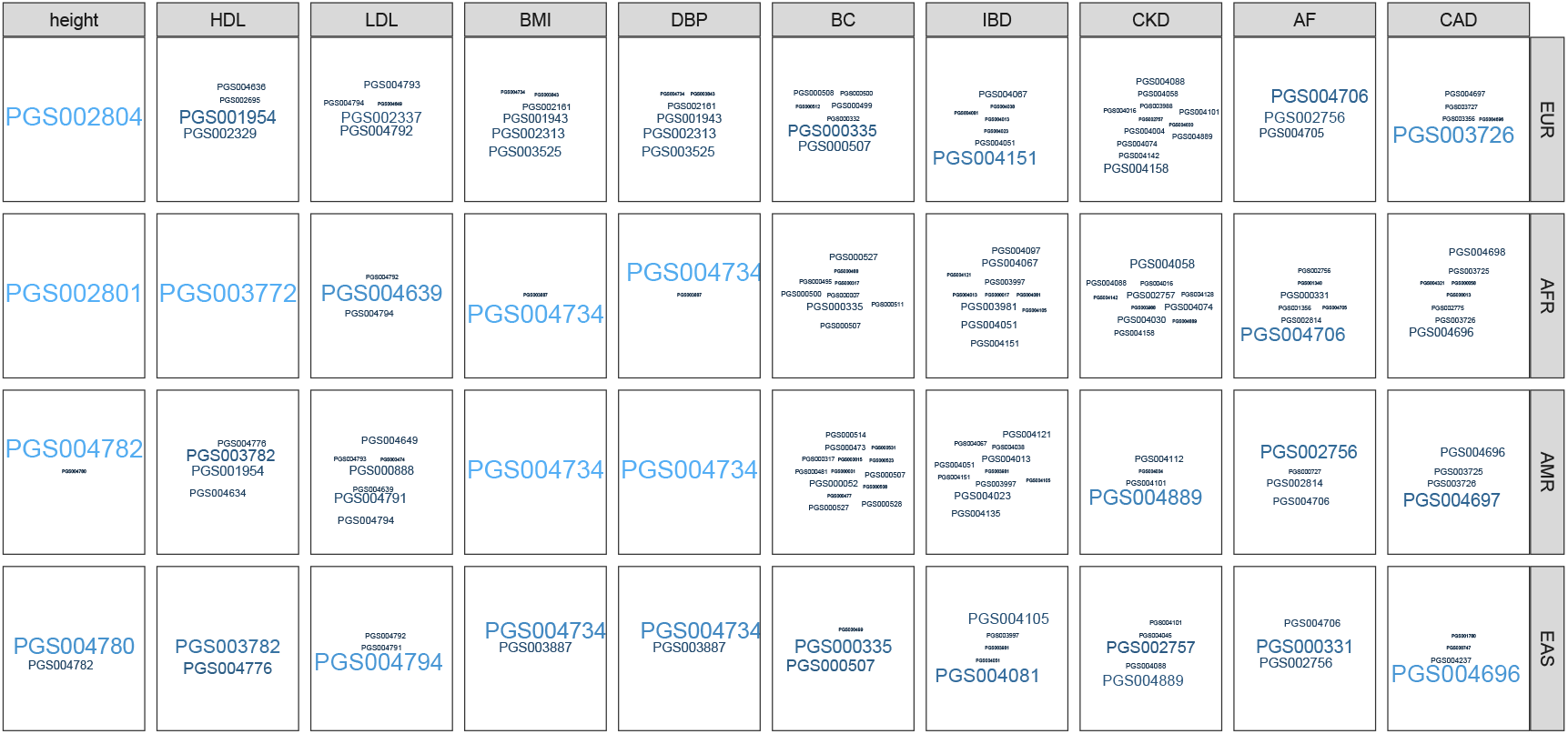
Word cloud visualization of the most frequently selected best PRS model across 100 repeated random samples (N = 10,000) for different traits and ancestry populations. The text size is proportional to the frequency with which each PRS model was identified as the best across the 100 draws.

Figure 4 demonstrates that for traits, such as height, the best PRS model is relatively consistent within each ancestral group, but varies across ancestry populations. The metadata of these PRS models reveals they are predominantly trained using GWAS studies based on the corresponding ancestry populations. In contrast for other traits, like LDL and BC, the best PRS model varies widely within and across ancestral groups, highlighting the poor generalizability and transferability of these trait-specific PRS models (Liu et al., 2021; Wang et al., 2022).

Although the weights in UNSemblePRS can be easily recalibrated for a new population, we also evaluate their transferability by applying the pre-learned UNSemblePRS weights directly to a new population, which has different context features, such as sex, age, and genetic ancestry, from the target cohort used for deriving the weights. Specifically, we train the best model and determine the weights for UNSemblePRS using a dataset of 10000 samples randomly selected from the AoU data. We then evaluate the model’s performance on an independent population drawn from the remaining AoU database, which exhibits a different distribution of sex, age or genetic ancestry. Transferability is assessed by comparing the relative performance changes between the independent samples and the target cohort.

Figure 5 presents the relative changes in predictive performance (measured by *R*^2^ for continuous traits and AUC for binary traits) averaged over all the considered traits across each context-specific population. Regarding the impact of genetic ancestry on the PRS prediction, performance changes are modest (*≤* 5%) on average in EUR population for both methods since the original target cohort used to learn the weights and select the best model is dominated by the EUR population. Notably, the predictive performance in UNSemblePRS tends to decrease less than that of the best model, with a decline of 1.8% for UNSemblePRS compared to 3.5% for the best model. The result indicates that UNSemblePRS is more generalizable to individuals who share the same ancestry origin but are not included in the target cohort (i.e., the independent samples from the EUR population). When the independent samples exclusively consist of the AFR, AMR, or EAS populations, the predictive performances show more pronounced declines. Specifically, the performance drop of the best model when transporting from the original cohort to the non-EUR populations ranges from 15.0% *−* 20.0%, while UNSemblePRS shows a smaller decline of 12.2%, with 15.9% for the AFR population, 8.5% for the AMR population, and 12.2% for the EAS population, respectively.

**Figure 5:**
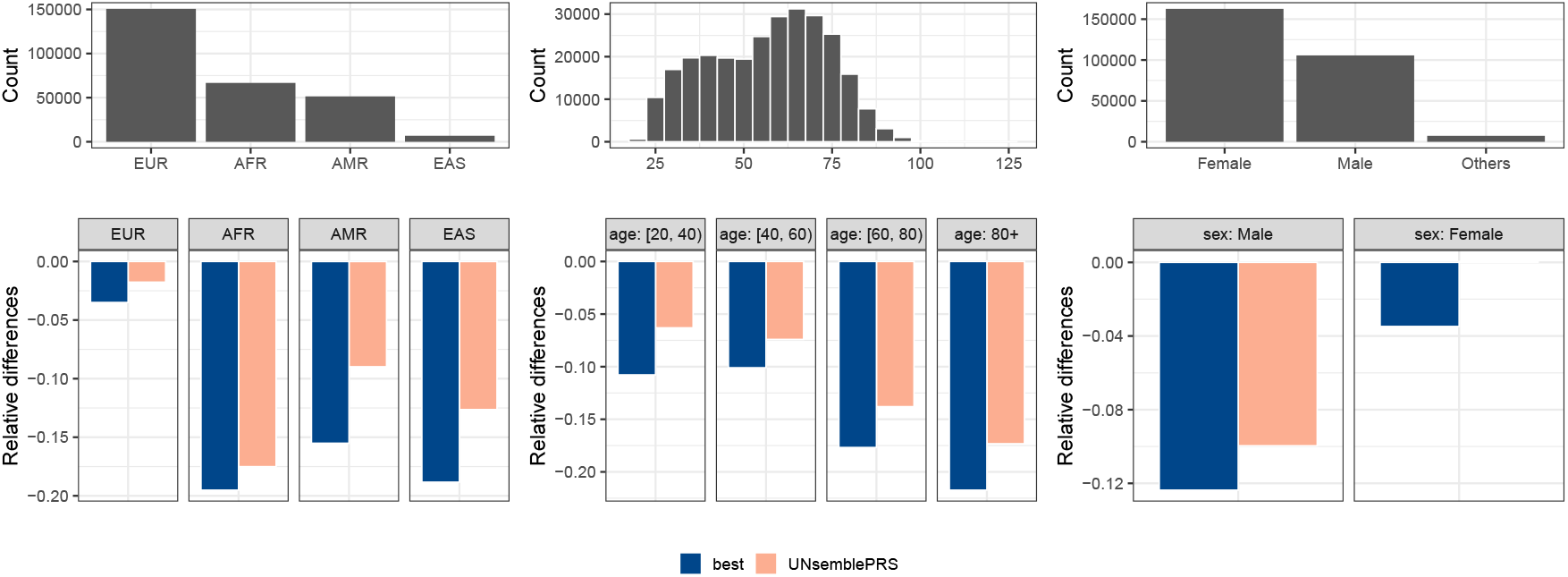
Relative changes in predictive performance of the best single PRS model and UNSemblePRS in independent samples compared to the target cohort, averaged over continuous and binary traits across 100 random draws (N = 10,000) for each context-specific population. The distributions of context-specific features for the target cohort are shown in the upper panel.

The performance drops when transporting from the original cohort to individuals aged between 20 and 60 have a decrease in PRS accuracy of 6.1% for UNSemblePRS compared to 10.4% for the best model. For those over 80, the decrease is 17.7% for UNSemblePRS compared to 21.7% for the best model. This disparity might be attributed to the large proportion of individuals between ages 20 and 60 (*≥* 50%) and the small proportion over 80 (*≤* 7%) in the target cohort, which leads to poor transferability for the senior population. Furthermore, UNSemblePRS shows improved transferability compared to the best model across sex-specific populations, with an average decline of 5% for UNSemblePRS compared to 8% for the best model. Overall, UNSemblePRS demonstrates smaller relative changes across both sex, age, and genetic ancestry when averaging over all traits, compared to the best model. This indicates that UNSemblePRS is less sensitive to shifts in contextual features in new independent samples.

#### Sparsity in UNSemblePRS helps to increase the predictive performance

The sparsity threshold in UNSemblePRS mitigates the impact of low-quality or corrupted models on the final aggregated PRS, using a data-driven approach. The sparsity level determines the proportion of models excluded by assigning their weights to 0 after the initial weights are derived from kernel PCA. The choice of the sparsity level can thus influence the performance of UNSemblePRS, and an optimal level may depend on prior knowledge about the candidate PRS models.

We compare the proposed UNSemblePRS methods with different sparsity levels in Figure 6, which illustrates the predictive performance across various levels of sparsity and ancestry populations, averaged over all the considered traits. Since UNSemblePRS utilizes kernel mapping for the relationship between PRS models, compared to PCA, it yields superior predictive performance for all the sparsity levels considered, except for the AFR population where low sparsity yields slightly worse performance than PCA. As the sparsity level increases, we see increased performance across all ancestry populations, as shown in Figure 6. This demonstrates that promoting sparsity is beneficial for selecting useful PRS models and removing unwanted redundant information. However, the sparsity level should not be set too high, as it risks excluding informative models and reducing performance. When using the PGS catalog as the source of pre-trained PRS models, we recommend a default sparsity level of 50%.

**Figure 6:**
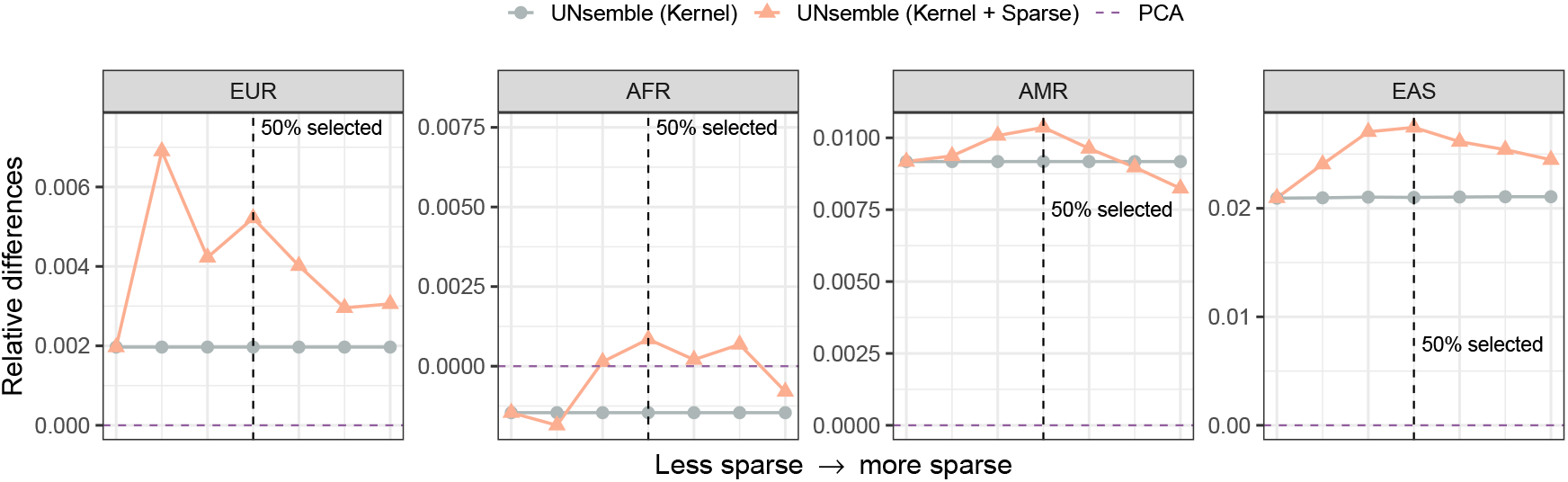
Changes in the relative performance of UNSemblePRS with and without sparsity, compared to PCA, stratified by ancestry populations and averaged across all traits, as the level of sparsity varies.

#### Scalability A key advantage of UNSemblePRS is its scalability. Leveraging sparse kernel PCA, weights can be computed in less than two seconds for a dataset with 10000 subjects and approximately 100 candidate models, enabling real-time implementation. For larger target sample sizes, UNSemblePRS supports distributed computing. This approach, partitions samples into random batches, computes kernel matrices within each batch, and combines the results. This strategy achieves runtime comparable to processing a single batch. Additionally, distributed computing facilitates the integration of data across multiple biobanks or institutions. This is particularly valuable when working with underrepresented populations, where combining data from multiple institutions can improve weight estimation. The distributed strategy allows for sharing only a *p × p* kernel matrix across sites, enabling joint learning of UNSemblePRS weights while maintaining data privacy. Details for the distributed algorithm are provided in the Online Method

## 3 Discussion

In response to the growing emphasis on open science and open-source machine learning and the increasing availability of pre-trained PRS models, we introduce UNSemblePRS, an efficient method for aggregating pre-trained PRS models to deliver robust and reliable performance in target cohorts in real time. By leveraging the numerous PRS models available in databases such as the PGS Catalog, UNSemblePRS avoids the complexity associated with calibrating or retraining models for target populations, which can often be computationally intensive and may be infeasible for some underrepresented populations. Using data from the AoU Research Hub, we demonstrate that UNSemblePRS achieves performance comparable to the best PRS model across diverse ancestry populations and traits. Additionally, it shows strong consistency between the weights assigned to each candidate model and their predictive performance. When applied to a new population, UNSemblePRS exhibits better transferability than the best model selected based on observed traits. Recalibrating the weights further enhances its predictive performance.

Despite the robustness of UNSemblePRS, performance gaps persist across ancestral groups, with predictive performance in non-EUR populations generally lower than in EUR populations. Even when comparing the best-performing model for each ancestral population, significant gaps remain for most traits. These disparities are rooted in the underrepresentation of non-EUR populations in genetic studies (Ju et al., 2022). In the PGS catalog, the majority of models are still trained on EUR GWAS or EUR-dominated GWAS due to the lack of genetic data from populations of non-European ancestry, leading to lower-quality models for non-EUR populations (Ding et al., 2022; Hou et al., 2024). As UNSemblePRS relies on the diversity and quality of its candidate set of pre-trained models, its performance in non-EUR populations is currently suboptimal. However, we anticipate that as recognition of these disparities grows and more diverse samples are collected, researchers will develop higher-quality models for underrepresented populations in the near future. UNSemblePRS will then play a critical role in leveraging the strengths of individual models to achieve improved performance in real-world applications.

Moving forward, the integration of pre-trained PRS models could benefit from incorporating external information about the reliability or similarity between the training data of the PRS models and the target population. For instance, the PGS catalog provides details on the ancestral composition of GWAS training and validation samples, as well as the methods and datasets used to develop the PRS models (Lambert et al., 2021, 2024a). While this information is not completely reported for all models, it could be utilized to guide the learning of optimal weights. However, striking the right balance between leveraging prior information and relying on purely data-driven weights will be critical. Users may need to carefully calibrate this balance, as the optimal approach could vary across different traits and populations.

## Data Availability

The code to implement UNSemblePRS is available on GitHub, https://github.com/biostat-duan-lab/UNSemblePRS and data from All of Us research Hub are available for registered users from https://www.researchallofus.org/data-tools/workbench/.

## Acknowledgement

We gratefully acknowledge All of Us participants for their contributions, without whom this research would not have been possible. We also thank the National Institutes of Health’s All of Us Research Program for making available the participant data examined in this study. This work was supported by the National Institutes of Health (R01MH137218 and R01GM148494).

## Data and Code Availability

The code to implement UNSemblePRS is available on GitHub, https://github.com/biostat-duan-lab/UNSemblePRS. This study used data from the All of Us Research Program, accessed through the All of Us Researcher Workbench https://researchallofus.org/. The dataset used in this study is from the Controlled Tier Dataset v7 (2022Q4R9) release. Data are available to approved researchers through the All of Us Researcher Workbench following completion of the required registration and training. Due to privacy and confidentiality policies, individual-level data cannot be shared directly. Researchers interested in accessing the All of Us dataset may apply for access through the All of Us Research Program.

## A Online Supplementary Material

### Online method for UNSemblePRS

Our unsupervised Polygenic Risk Score (PRS) ensemble method is grounded in the spiked covariance model (Johnstone, 2001), which has been extensively utilized across a range of applications (Lobach and Detmer, 2007; Salmon et al., 2014; Cao et al., 2020). In this model, let *X*_*n×p*_ represent the standardized matrix consisting of *p* PRS scores for *n* individuals with respect to a single phenotype, where each column has a mean of zero and a unit length. Standardizing multiple PRSs helps to mitigate variations in model scales and ensures comparability across models. We assume that the covariance matrix of the *p* PRS models, denoted by Σ_*p×p*_ follows Σ_*p×p*_ = Ω_*p×p*_ + *σ*^2^*I*_*p×p*_ where Ω_*p×p*_ = *vv*^T^ and *v* is a unit vector representing relatively each model’s performance, and *σ*^2^*I*_*p×p*_ represents the noise induced by sampling and other prediction errors. The unit vector *v* is referred to as the spike, which indicates the strength of the predictive signal for each PRS model, and therefore reflects each model’s importance.

Our goal is to recover the vector *v* and use it as the aggregation weight for the unsupervised ensemble learning. The estimation of *v* is equivalent to estimating the eigenvector of the sample covariance matrix Σ_*p×p*_. However, the sample covariance Σ_*p×p*_ only captures the linear relationships among these PRS scores. To generalize, we apply a kernel function Φ to map the score matrix *X*_*n×p*_ into a non-linear space *F*_*p×p*_, where nonlinear relationships among the PRS scores can be explored. In UNSemblePRS, the default kernel function Φ is chosen to be the polynomial kernel with a degree of 3. Specifically, for PRS scores *X*_*·*,*k*_ and *X*_*·*,*k*_^*′*^ predicted by the *k*-th and *k*^*′*^-th PRS models, we measure their relationships by computing 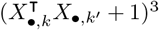for the (*k, k*^*′*^)-th entry of the feature space *F*_*p×p*_; other kernel functions are available in the R package `kernlab`.

Next, we employ the sparse PCA algorithm from Erichson et al. (2020) to remove redundant signals from the kernel space. More specifically, the rank-one sparse PCA algorithm is applied to the mapped kernel spaces, minimizing the following objective function:

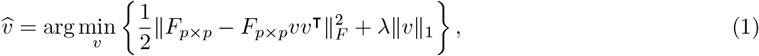

where *λ* is the tuning parameter, and ∥*v*∥_1_ is the *𝓁*_1_ norm for the vector *v* to promote sparsity. Finally, the final aggregated PRS score is derived by combining the standardized PRS matrix *X*_*n×p*_ according to the aggregation weights *v*; see Algorithm 1 for the detailed procedures.

### UNSemblePRS for distributed learning

The UNSemblePRS is particularly suited for a distributed learning framework. By computing the correlation matrix Σ^(*s*)^ for each site-specific PRS score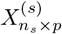as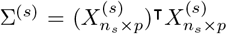, we can obtain the distributed correlation matrix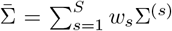 for the central server, where *w*_*s*_ are user-specified site weights that reflect prior information about the data quality at each site. Next, we extract the distributed feature space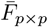 by applying the polynomial kernel to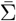. Finally, the distributed aggregation weights are obtained by applying the rank-one sparse PCA algorithm to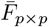. Distributed learning for other kernel functions should be discussed on a case-by-case basis and is omitted here. This process refines the ensemble learning approach while preserving privacy concerns and offers a more accurate polygenic prediction for a more complex target cohort; see Algorithm 2 for the complete implementation.

#### Algorithm 1

UNSemblePRS Method for PRS Ensemble Learning

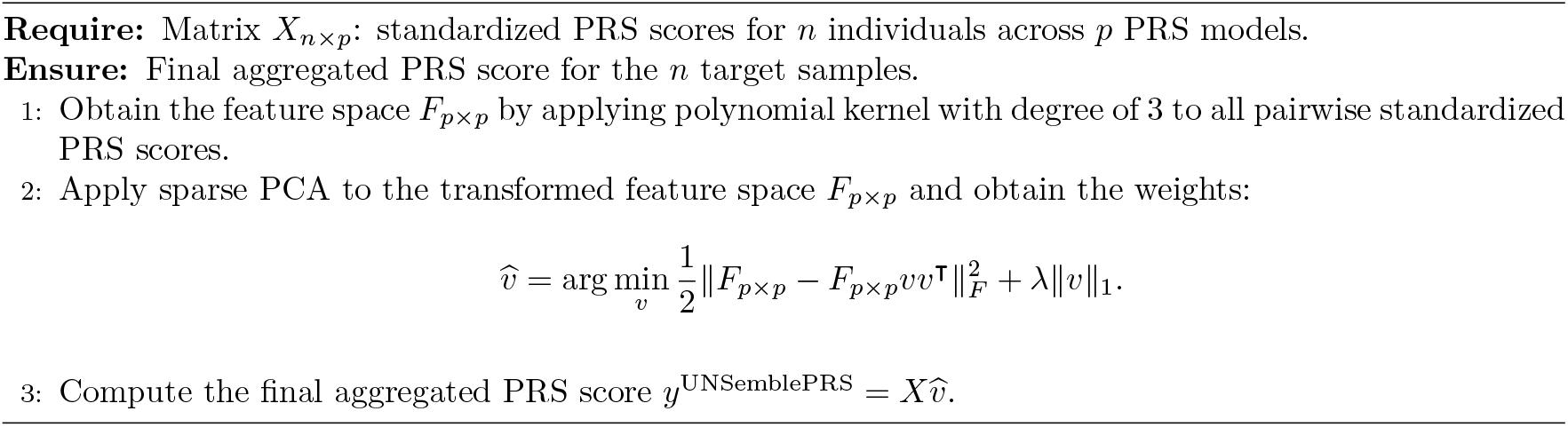

#### Algorithm 2

Distributed UNSemblePRS Method for PRS Ensemble Learning

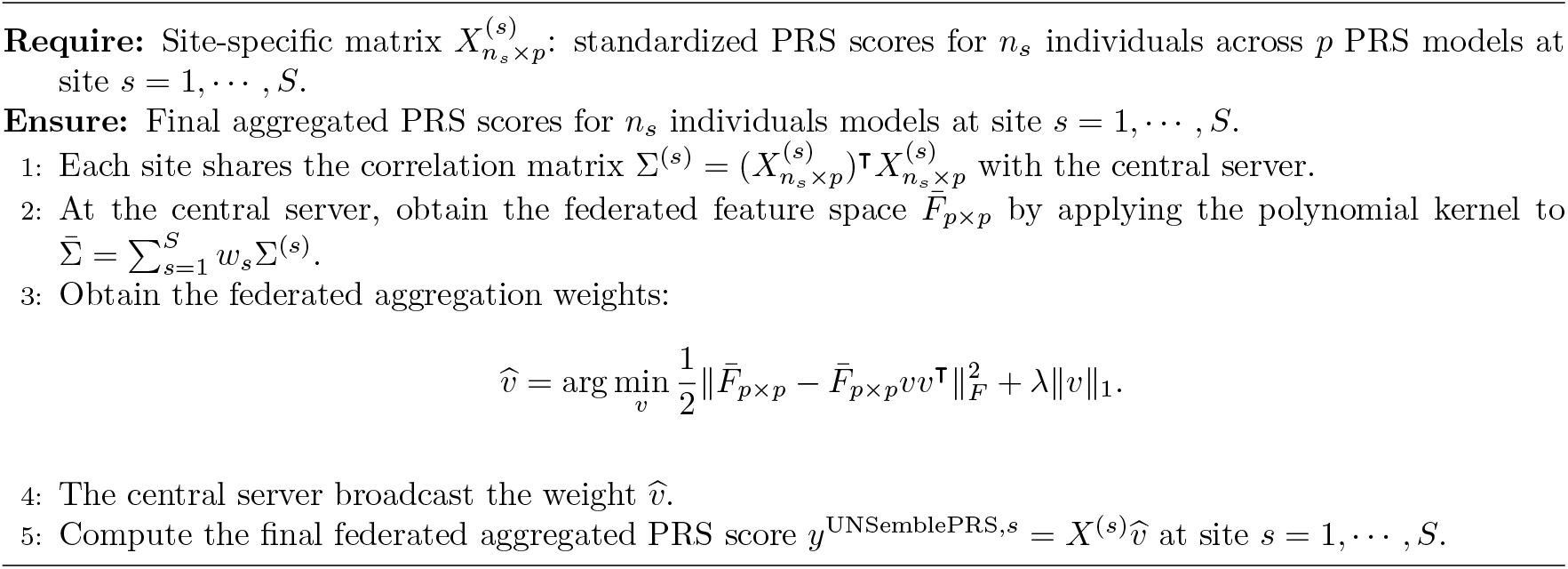

### AoU Data extraction and curation

The All of Us (AoU) Research Program is a population-based, longitudinal initiative designed to recruit at least one million participants from across the United States to advance precision medicine (All of Us Research Program, 2019). Ongoing enrollment captures extensive health-related data—ranging from genomic data and electronic health records to lifestyle factors and environmental exposures—to elucidate how these elements collectively influence health and disease. Our analyses leverage the latest All of Us Controlled Tier Dataset v7 (2022Q4R9). The program operates under a centralized institutional review board, and all participants provide informed consent. The present study is a secondary analysis of de-identified AoU participants, approved as nonhuman participants research by the AoU Research Program Resource Access Board, with a waiver of the informed consent from the participants.

#### Genotyping and quality control

Genotyping and primary quality control (QC) are performed by the centralized data processing team of the AoU Research Program, summarized previously (All of Us Research Program, 2024). The AoU microarray genotyping dataset consists of 1.8 million single nucleotide variants (SNVs) genotyped for 312925 participants, mapped to the hg38/GRCh38 reference genome. QC ensures that samples are sex-concordant (i.e., genotype-based self-reported sex match at birth), have a missing call rate below 2%, and contamination rates under 3%. Genetic ancestry and relatedness are determined for the participants with genome-wide short-read sequencing data.

#### Polygenic scoring

For PRS generation, we use genotyped SNPs in the 1000 Genomes Project Phase 3 because this reference panel offers comprehensive global representation, compatibility with the AoU dataset’s genetic architecture, and reliable imputation accuracy, making it well-suited for ancestry-aware polygenic score analyses.

#### Ancestry annotation

Ancestry groups are defined based on the Human Genome Diversity Project and 1000 Genomes reference panels, including African/African American (AFR), Admixed/ Latino/ Native American (AMR), East Asian (EAS), European (EUR), Middle Eastern (MID), South Asian (SAS), and Other/Multi (OTH, representing individuals not belonging to a single ancestry or with balanced admixture). For the remaining 285269 individuals with genome-wide genotyping data, ancestry and relatedness are assigned using a K-means clustering algorithm (*k* = 5) applied to 16 genetic principal components (PCs), as described by the AoU Genomics Initiative. Within each predicted ancestry population, PCA is performed using PC-AiR in the R GENESIS package to account for genetic sub-structuring while controlling for relatedness. Phasing and imputation are conducted with ShapeIT (v5.0) and minimac (v4-1.0.2), using the 1000 Genomes Project Phase 3 as a reference panel. Post-QC genotype data includes bi-allelic SNPs with a missing rate below 2%, Hardy-Weinberg equilibrium p-value less than 1 *×* 10^*−*5^, minor allele frequency above 0.1%, and imputation quality scores of at least 0.8 for each population.

#### Extraction of traits and covariates

The considered traits for the AoU database are summarized in Table A1. Continuous traits are curated as the average of the most recent five measurements, or fewer if less than five measurements are available. Phenotype labels for the considered binary traits are determined as cases when individuals have been diagnosed based on ICD and SNOMED criteria, or a documented disease history for the target trait. Age for each individual is defined as the difference between the current year, 2023, and the individual’s birth year.

**Table A1:** Summary of considered traits, number of models, and sample sizes of the AoU data.

|  | Phenotype | EFO | # PRS | # Participants |
| --- | --- | --- | --- | --- |
| Continuous | Body Mass Index | EFO_0004340 | 93 | 268834 |
|  | Height | EFO_0004339 | 85 | 269570 |
|  | High-Density Lipoprotein | EFO_0004612 | 90 | 113180 |
|  | Low-Density Lipoprotein | EFO_0004611 | 104 | 85890 |
|  | Diastolic Blood Pressure | EFO_0006336 | 51 | 222367 |
| Binary | Breast Cancer | EFO_0000305 | 135 | 223106 |
|  | Inflammatory Bowel Disease | EFO_0003767 | 20 | 223106 |
|  | Chronic Kidney Disease | EFO_0003884 | 20 | 223106 |
|  | Atrial Fibrillation | EFO_0000275 | 37 | 71437 |
|  | Coronary Artery Disease | EFO_0001645 | 67 | 71437 |

### Model evaluation for quantitative and binary traits

We randomly draw 10000 individuals from the AoU data who have both genotypic data and observed trait of interest as the target cohort. The number of participants with observed phenotypes for each trait is presented in Table A1. A typical study involving PRS aims to test the association between PRS and a target trait. This association is evaluated using standard metrics for association testing or goodness-of-fit assessment. For continuous traits, once covariates such as age, sex, and genetic principal components are accounted for in the model, we report the incremental phenotypic variance explained, denoted by *R*^2^. This measure represents the increase in *R*^2^ due to the inclusion of the PRS in the regression model, thereby isolating the unique explanatory power of the PRS itself. When dealing with binary traits, a common practice is to assess the model’s discriminative ability through the area under the Receiver Operating Characteristic curve (AUC), adjusted for covariates to ensure the contribution of PRS is accurately reflected (Zhang et al., 2023).

Each PRS method is applied to the target cohort stratified by sex, age, and ancestry annotation. The prediction accuracy is assessed across 100 repeated draws of target cohorts (Figure 2, Figures A1 and A2). The trait BC is excluded from the analysis for the stratified evaluation based on sex. The best model is selected from the candidate PRS scores with the largest association with the target trait in the given stratified population. The average of PRSs, weighted by the size of the relevant GWAS studies, and the PRS derived from the largest GWAS study are based on a subset of the candidate PRS scores with metadata available at the PGS Catalog. The aggregation weights for the PCA and UNSemblePRS methods are obtained based on the stratified population. The sparsity level for the UNSemblePRS is set to 50% by default using candidate PRS scores from the PGS Catalog.

Furthermore, for each trait considered, both predictive metrics and aggregation weights of candidate PRS models are standardized to have zero mean and unit variance. The scaled predictive metrics, measured by *R*^2^ for continuous traits and adjusted AUC for binary traits, are then compared with the scaled aggregation weights derived from UNSemblePRS and standard PCA analysis in scatter plots (Figure 3). Each point represents a PRS model, and the curves are fitted by linear regression models for these two methods, respectively. The concordance between the actual model performance and the assigned weights is computed as the *R*^2^ of the fitted linear regression models.

**Figure A1:**
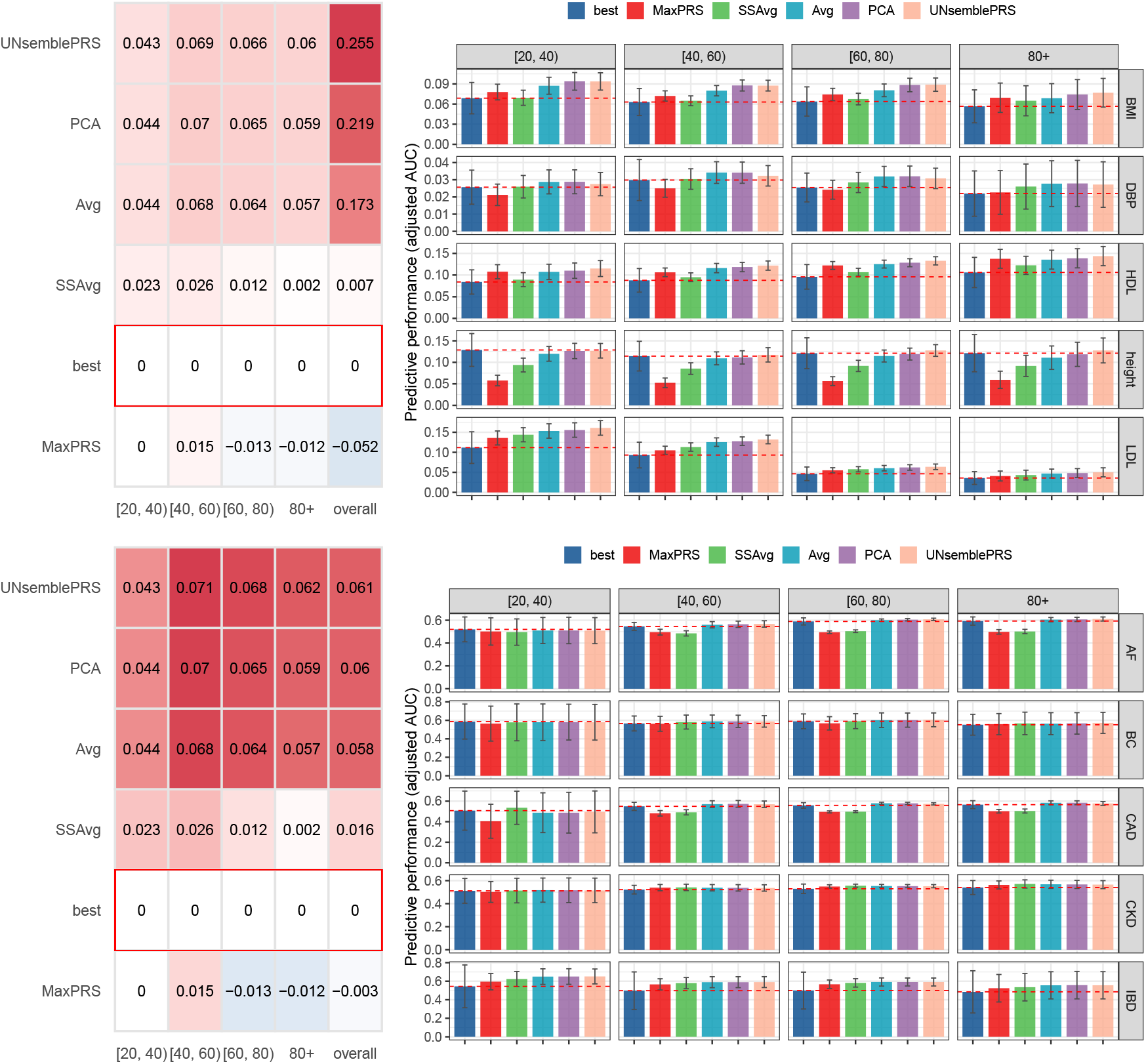
**(Left)** Relative differences in predictive performance compared to the best single PRS model, evaluated across 100 random samples (N = 10,000) and averaged over continuous and binary traits in the overall target cohort and stratified by age groups. **(Right)** Predictive performance for continuous and binary traits in the overall target cohort and across age groups. Error bars represent*±*1 standard deviation computed from the random samples. Dashed lines denote the performance of the best single PRS model.

**Figure A2:**
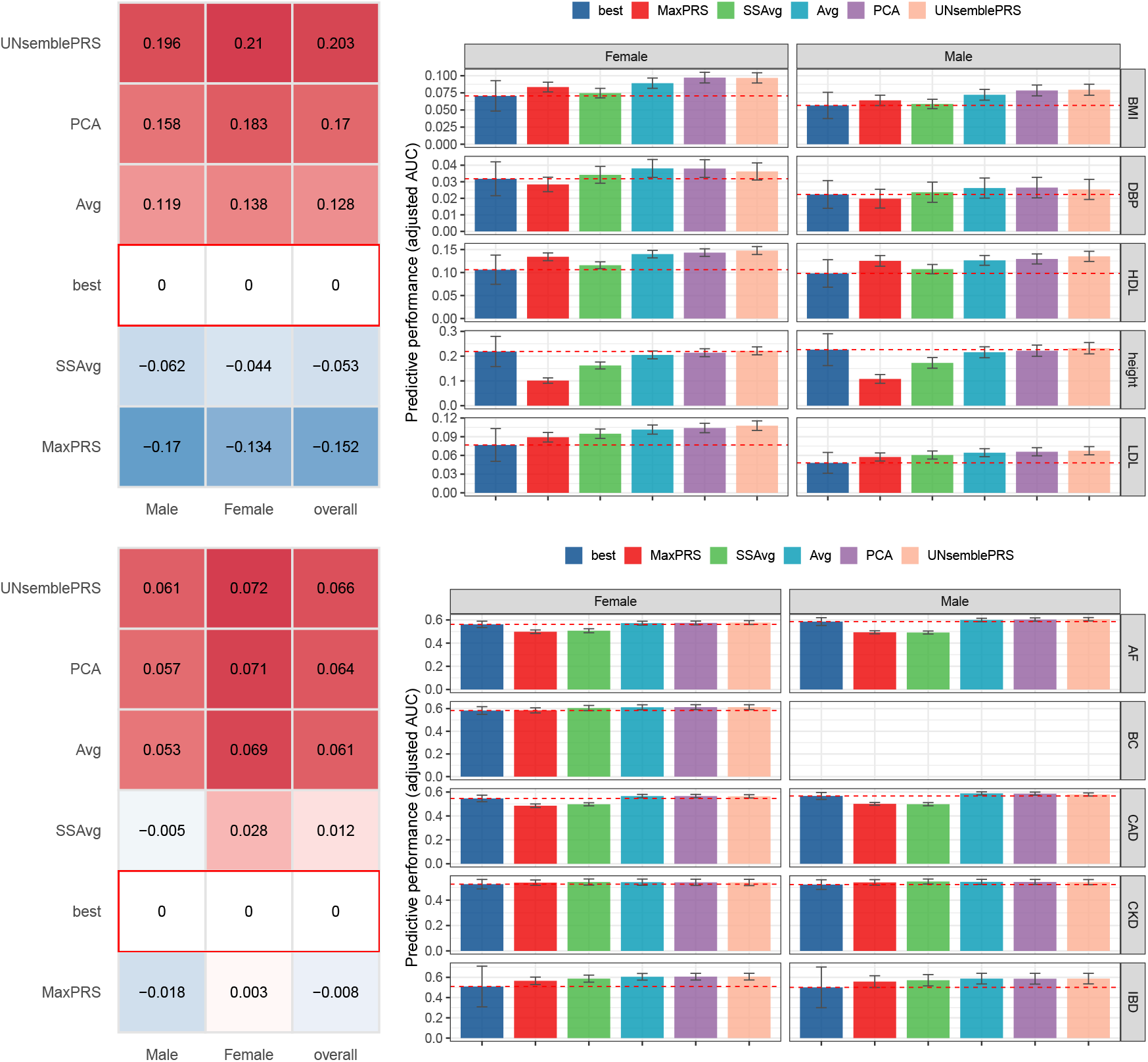
**(Left)** Relative differences in predictive performance compared to the best single PRS model, evaluated across 100 random samples (N = 10,000) and averaged over continuous and binary traits in the overall target cohort and stratified by sex groups. **(Right)** Predictive performance for continuous and binary traits in the overall target cohort and across sex groups. Error bars represent *±*1 standard deviation computed from the random samples. Dashed lines denote the performance of the best single PRS model.

### Assessing model transferability

To assess the transferability of the UNSemblePRS, we apply the prelearned UNSemblePRS weights from the target cohort to independent samples that consist exclusively of one ancestry population. Specifically, for each randomly sampled target cohort, we randomly draw independent samples from the remaining individuals in the AoU database, ensuring the independent samples have the same sample size as the target cohort and consist exclusively of a single ancestry population (i.e., EUR, AFR, AMR, or EAS) or include individuals from a specific age ranges (i.e., [20, 40), [40, 60), [60, 80), and 80+) or sex groups (male or female). The changes in predictive performance relative to the target cohorts are computed for the UNSemblePRS and the best single PRS models, as presented in Figure 5.

### Sensitivity analysis on the impact of varying sparsity threshold

To examine the impact of sparsity level on the performance of UNSemblePRS, we assess the model’s predictive performance by applying the UNSemblePRS to the target cohorts under varying levels of the tuning parameter *λ*. Specifically, the proportion of selected candidate PRS models is ranging from 100% to 10%. The changes in predictive performance relative to the PCA analysis are computed for the UNSemblePRS with and without sparsity, as well as for the best single PRS model, as presented in Figure 6. This evaluation allows us to understand the sensitivity of the UNSemblePRS to different sparsity levels and how these levels impact the performance of ensemble learning for the considered traits.

### Sensitivity analysis on the impact of kernel function

To examine the impact of various kernel functions on the performance of UNSemblePRS, we evaluate the model’s predictive performance using three distinct kernel functions: 1) a polynomial function of degree 3, 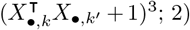 a polynomial function of degree 4, 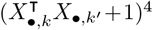 and 3) a Gaussian radial basis kernel, exp(−∥*X*_*·*,*k*_ *−X*_*·*,*k*_^*′*^ ∥). Each UNSemblePRS model is assessed on the replicated ancestry-specific target cohorts. The results are presented in Figure A3. It can be observed that UNSemblePRS achieves prediction accuracy comparable to the best single PRS model across different ancestry populations, regardless of the chosen kernel function. On average, the predictive performance for the continuous traits is 0.083 for the polynomial kernel and 0.080 for the Gaussian radial basis kernel, while the predictive performance for the binary traits is 0.568 for the polynomial kernel and 0.561 for the Gaussian radial basis kernel. This suggests that the model can be robust to kernel choice, provided that some level of flexible nonlinear relationships is captured. In practice, the selection of a specific kernel function should be made on a case-by-case basis, guided by domain knowledge.

**Figure A3:**
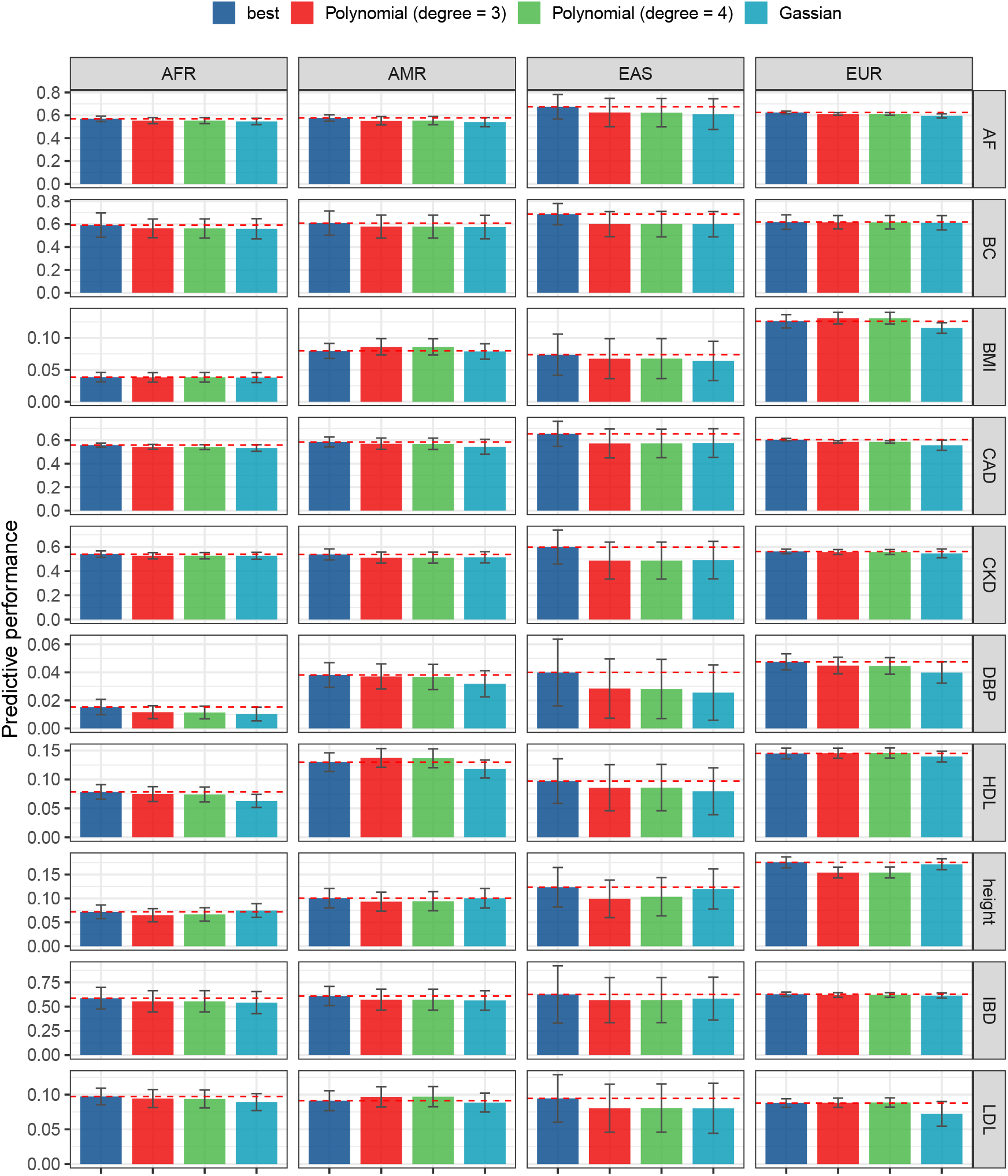
The predictive performances of the UNSemblePRS with different kernel functions for continuous and binary traits on the overall target cohort and separately on each ancestry population.

